# Treatment pathways of non-small cells lung cancer patients in the Czech Republic: an insight from administrative claims data

**DOI:** 10.1101/2025.07.25.25332126

**Authors:** Aleš Tichopád, Gleb Donin, Vratislav Sedlák, Marian Rybář, Martin Rožánek, Karla Mothejlová, Vladimír Koblížek, Pavel Turčáni, Milan Sova, Ladislav Dušek, Zuzana Bielčiková

## Abstract

**Introduction:** A patient pathway is an evidence-based tool that details the phases of care with the aim of increasing the effectiveness and efficiency of patient care.

**Methods:** This was a longitudinal, historical descriptive cohort study spanning from 2017 to 2022. The index date was determined by the first bronchoscopy (BX) followed by histopathological (HP) examination, alongside the presence of the ICD-10 diagnosis code C34. Incident patients aged ≥18 years were included if they had no prior malignancy reported in the data. Pharmacotherapies (PHT), including chemotherapy (CT), precision therapy (IOTT), as well as surgery (SX) and radiotherapy (RT), were investigated concerning overall survival. Additionally, the presence of a reported multidisciplinary team (MDT) and treatment at a Complex Oncological Center (COC) with high-load experience was considered.

**Results:** By using administrative claim data we analyzed the pathways of 16,783 NSCLC patients with the aim to provide a clear depiction of the current state of care. Less than 40% (38.4%) of patients had MDT reported within a median of 21 days. Of the 12,803 treated patients,, 47.42% received CT, 27.56% underwent SX, 1.4% underwent RT, and 5.32% IOTT. Early initiation of treatment within 4 weeks from BX was identified in only 20% of SX patients, 24% of patients treated with CT, and 26% of RT patients. The centralization of care in COCs primarily concerned SX and CT_NEO, while 24% of patients indicated to CT were treated elsewhere. The OS rate of those treated was 31% in median, 64.3% in SX patients, and 14% in those treated with CT. The prognosis of patients was positively affected by COCs.

**Conclusions:** We developed a methodology for administrative data processing which can be implemented as a technical infrastructure to fulfill the organization and quality evaluation of cancer care in the Czech Republic.

**KEY MESSAGES:**

- There is limited evidence regarding the effects of patient pathways used in oncological care. However, there is a need for the integration of innovative approaches to advance cancer control.
- We have developed a methodology for processing administrative data that can be implemented alongside the Czech National Cancer Registry to create a comprehensive dataset including histopathological characteristics, disease stages, and reimbursement care data.
- A comprehensive Information and Communications Technology model, integrating multiple data sources, has been prepared to facilitate the organization and quality evaluation of cancer care in the Czech Republic.

## INTRODUCTION

Lung cancer stands as the most fatal cancer globally [1], with approximately 85% of patients falling under the category of non-small cell lung cancer (NSCLC), which encompasses various histological subtypes, most commonly lung adenocarcinoma and lung squamous cell carcinoma [2]. Accurate determination of the specific lung cancer type is vital for guiding subsequent treatment decisions. Recent comprehensive evaluations of lung cancer survival rates in Europe reveal a 39% one-year survival rate and a 13% five-year survival rate, with significant disparities across regions [3]. In the Czech Republic, the five-year relative survival rate for treated lung cancer patients has doubled from 9.6% to 19.3% in the past 20 years [4]. Advancements in immunotherapy, particularly with PD-1/PD-L1 inhibitors, offer promise for improved treatment outcomes [5–8].

Early identification of the disease is crucial, as surgical resection of NSCLC at an early stage offers favorable prognoses [9], with reported five-year survival rates of up to 70% for small, localized (stage I) tumors [10]. However, population-based data on lung cancer reveal that over 65% of new cases in the Czech Republic are diagnosed at clinical stage III and IV, with corresponding five-year stage-adjusted relative survival ratios below 15% and 5%, respectively [4]. Similar trends are expected globally [11], contributing to the elevated mortality rate from lung cancer due to the diagnosis of metastatic disease in most patients [12].

Common pathways to diagnosis include GP-ordered imaging and lung specialist attendance without emergency hospital admission, with significant variations in time to diagnosis based on cancer stage [13]. Access to skilled practitioners who can successfully conduct bronchoscopy with sufficient tissue yield is a key milestone, particularly considering patient reluctance to undergo repeat examinations [14,15]. Additional tissue sampling or sample analysis at a remote specialty lab may also be necessary for molecular profiling of tumor mutations to guide the use of targeted therapies [16]. Utilizing a dedicated gatekeeper interventional pulmonology practice has been shown to decrease wait times from abnormal imaging to treatment initiation in newly diagnosed patients in the USA [17].

According to clinical guidelines, the assessment of operability and disease severity should occur within a multidisciplinary team (MDT), which plays a vital role in early care and is presumed to significantly impact clinical outcomes [18,19]. However, establishing a randomized controlled trial to investigate the effect of MDT remains challenging. On the other hand, there is evidence of the significance of high-volume oncological surgical care [20] and the necessity for centralization of thoracic surgery [21].

Since January 2022, lung cancer screening for high-risk individuals has been implemented in the Czech Republic as a nation-wide 5-year pilot study [22]. This potentially groundbreaking initiative has the potential to significantly alter the distribution of cancer stages diagnosed in individuals suspected of having lung cancer. To further inform health policy decisions, understanding the key milestones along the patient journey following a suspicious finding is crucial. Large electronic health records and administrative data have proven invaluable for understanding this subject [13, 23–25].

The Czech Republic’s membership in the Innovative Partnership for Action Against Cancer Joint Action (iPAAC) focuses on the creation of a comprehensive Information and Communication Technology (ICT) model [26]. This model aims to integrate multiple data sources into a functional national cancer care information system for the collection and publication of cancer care performance indicators. The National Health Information System (NHIS) serves as the foundation, providing representative data on healthcare providers, professionals, and consumed/reimbursed services in the Czech Republic. The National Registry of Reimbursed Health Services (NRRHS) is one of the most important components of the entire system, capable of overseeing all consumed healthcare services.

## METHODS

### Study design and data sources

This was a longitudinal, historical descriptive cohort study conducted using real-world administrative data from the insured Czech population. We utilized anonymized health administrative claim data covering the time period from 2017 to 2023, which provided de-identified information about patient interactions with the healthcare system, including age, sex, medical specialty, International Classification of Diseases 10th revision (ICD-10) diagnoses, and the date of death where reported. Information on tumor stage was missing from this analysis.

### Study period, index date and cohort selection

The study design and cohort selection are depicted in Figure 1. The overall study period extended from January 1st, 2017, to December 31st, 2023, overlapping with the available administrative data period for our study. The study cohort selection period ran from January 1st, 2017, until December 31st, 2022. Additionally, patients with less than 12 months of administrative data coverage were excluded (i.e., index date after December 31st, 2022) to facilitate the investigation of study outcomes during this period.

**Figure 1.**
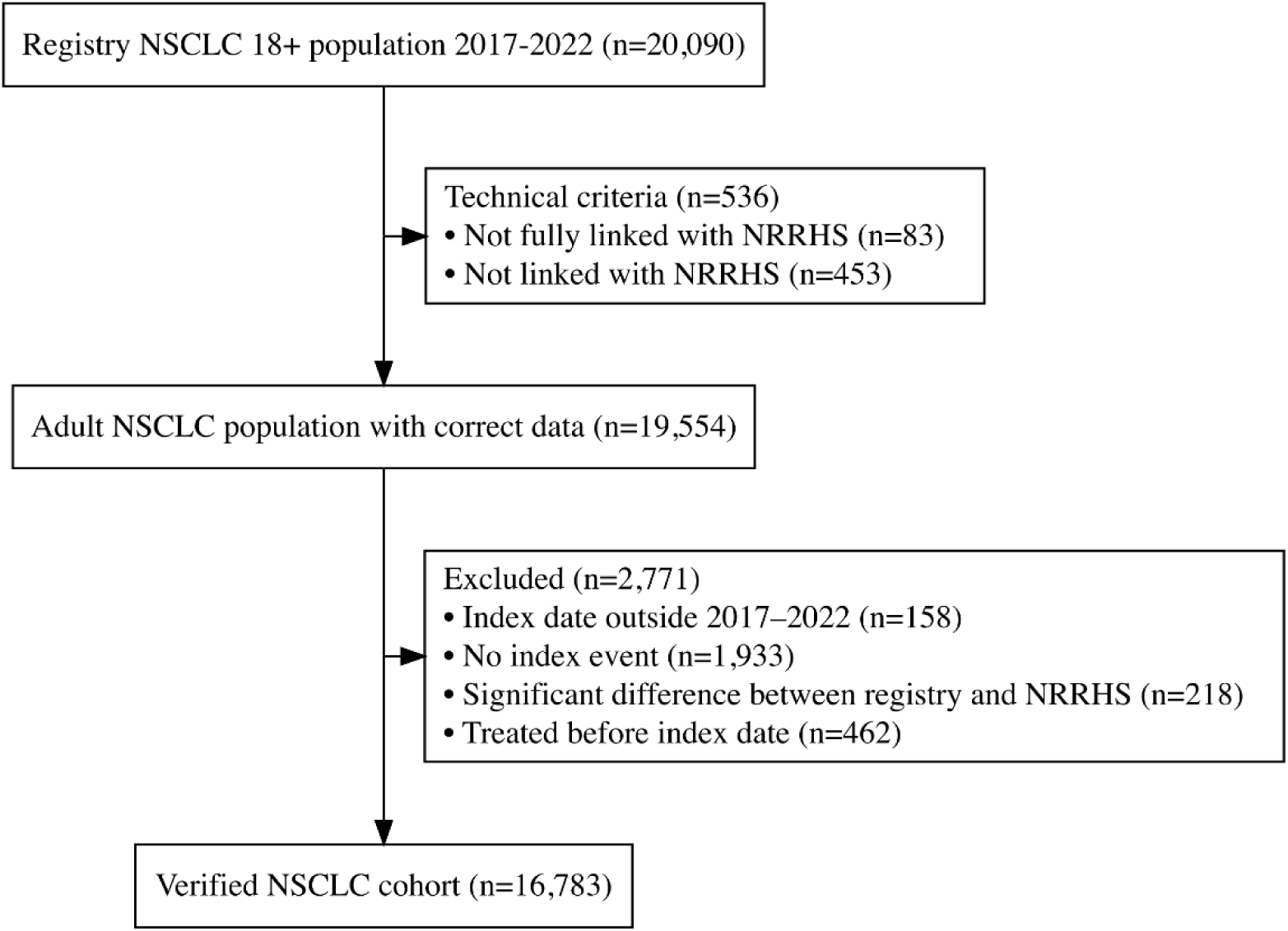
Overview of study design and patient cohort selection. Abbreviations: *ICD-10* International Classification of Diseases, 10^th^ revision; *C34* lung cancer

To establish the study cohort, the index date was defined as the date of the first bronchoscopy with lung biopsy (BX) followed by histopathological examination (HP) within the next 60 days, both occurring during the study period. Additionally, the ICD-10 diagnosis code C34 needed to be reported on at least one claim document of either the BX or the HP, or on another document within 180 days following the later of either the BX or the HP. The cohort selection was defined as those with an index date falling within the period from January 1st, 2017, to December 31st, 2022, and aged ≥18 years on the index date. Furthermore, patients were excluded if they had no single inpatient or outpatient claim of any type during the calendar year preceding the index date, or if they had a claim related to cancer surgery (SX), radiotherapy (RT), or pharmacotherapy (PHT) prior to the index date.

### Pharmacotherapies

Among PHT, in addition to the chemotherapy subgroup (PHT_CT), we further subdivided the precision therapy (PHT_IOTT) category. This included ATC codes for both immunotherapy (PHT_IO) and targeted therapy (PHT_TT). Furthermore, we defined neoadjuvant chemotherapy (PHT_CT_NEO) as any chemotherapy administered prior to SX and followed by SX within 6 months of its initiation.

### Follow-up period

The follow-up period was defined as the time from the index date to the first occurrence of any of the following events: (i) the end of the calendar year with the last patient record in administrative data; (ii) the end of study period (December 31st, 2023); (iii) death.

### Objectives and outcomes

The objective of the study was to describe the patient pathways to therapy for NSCLC patients in the Czech Republic and their survival during the period before the introduction and widespread use of preventive screening for lung cancer, i.e., prior to 2023.

### Statistical analysis

The statistical analysis was conducted using R for Windows. In addition to reporting absolute and relative frequencies and employing descriptive statistics for quantitative variables, we conducted a Kaplan-Meier analysis and estimated a log-rank as well as Cox proportional hazards model to study the centralized first-line treatment in COC and the involvement of a MDT review as factors associated with survival. These associations were studied separately for key treatment modalities such as SX, PHT, or RT, as each may more homogeneously represent a particular stage of the disease, thereby compensating for our lack of knowledge regarding tumor staging.

## RESULTS

### Study cohort

Our study cohort consisted of 16,783 patients, compromising 64% males (10,703) and 36% females (6,080). The mean age of the cohort at the index was 69 years (SD 9 years). Patients were evenly distributed over a span of four years, with the following annual distributions: 17% (2,881) in 2017, 17% (2,860) in 2018, 18% (3,074) in 2019, 16% (2,617) in 2020, 16% (2,724) in 2021 and 16% (2,627) in 2022. All 16,783 patients in the study cohort initially underwent BX with lung/bronchial biopsy.

The study details of the group of treated patients (12,803) are displayed in online supplemental Table S1. The median age and sex distribution were similar to those of the total patient population.

### Patient pathways

Patient pathways are described in Figure 2. Following the diagnosis (BX and HP) of NSCLC 22.89% of 16,783 patients died within 52 days in median without receiving treatment or evaluation by the MDT (Figure 2A). In total, 0.82% of patients were censored before receiving any treatment, indicating they were lost to follow-up or the study ended before their death could be recorded. Less than 40% (38.4%) of patients received MDT review within a median of 21 days. Conversely, 30.73% of patients underwent SX with a median time of 25 days, 50.37% received PTH with a median time of 21 days, and 18.9% underwent RT with a median time of 35 days post-MDT.

**Figure 2.**
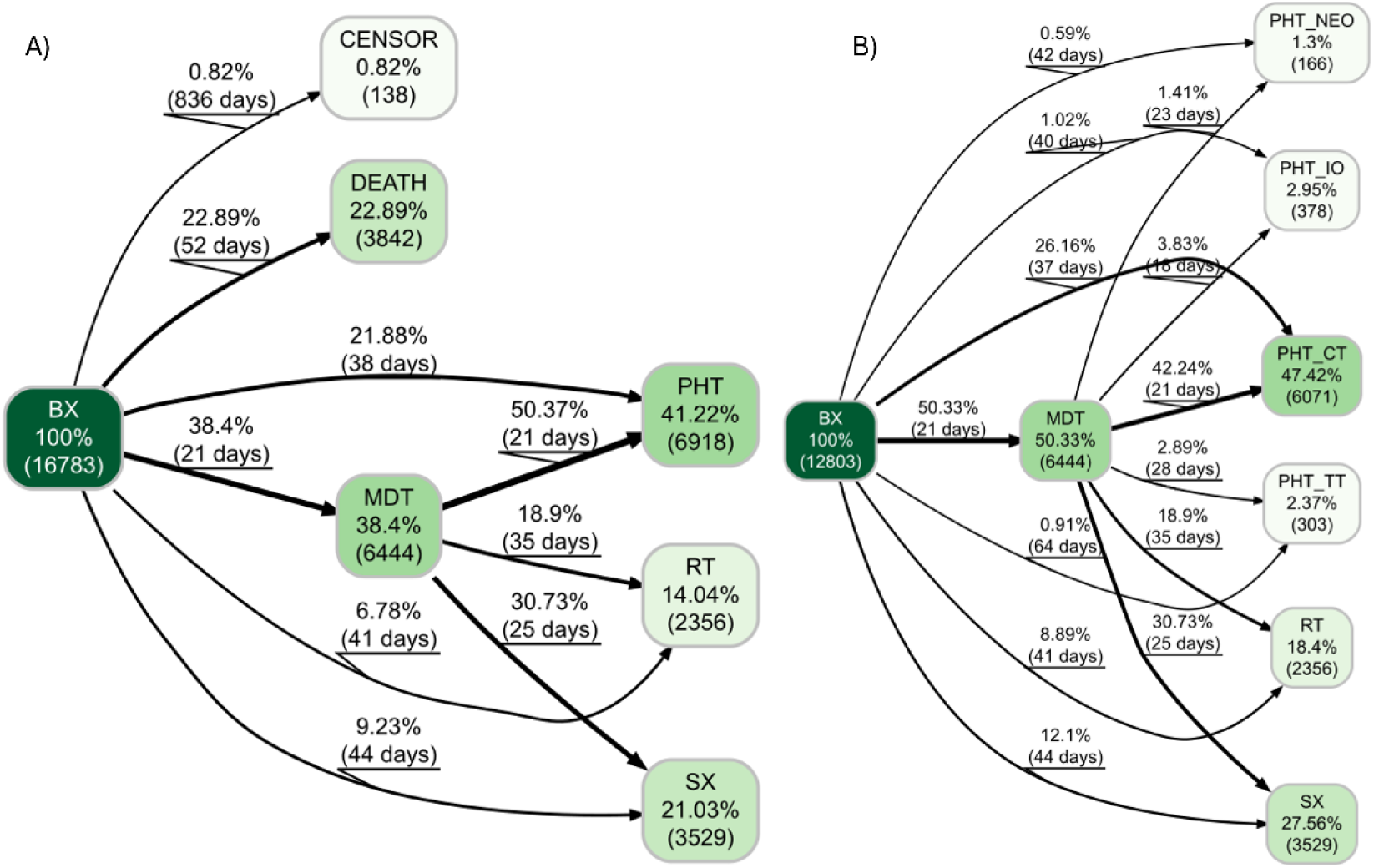
Patient pathway trajectories directly following the initial bronchoscopy in NSCLC patients. 1A: The pathways illustrate the progression of patients through various stages of the healthcare system after the initial bronchoscopy (BX). The numbers in the nodes correspond to the relative and absolute number of patients. The percentage on the edges represents the proportion of the population from the previous node. The number of days on the edges corresponds to the median difference between event onsets. 1B: This demonstrates a filtered cohort of patients who receive first-line treatment after BX. Abbreviations: *PHT* pharmacotherapy; *CT* chemotherapy; *CT_NEO* neoadjuvant chemotherapy; *IO* immunotherapy; *TT* targeted therapy; *BX* bronchoscopy; *RT* radiotherapy; *SX* surgery; *MDT* multidisciplinary team

Among the 12,803 of treated patients, the distribution of first-line therapy was as follows: 27.6% of patients underwent SX, 54.04% started PHT, and 18.4% received RT (Figure 2B). Additionally, 43.9% of operated patients and 55.2% of patients indicated for PHT_CT were not discussed in the MDT.

### Time to treatment

The median treatment initiation time was 46 days (see online supplemental Table S1). Patients treated with PHT_CT had the shortest median at 42 days (range 28-64), possibly due to less demanding diagnostics processes, followed by PHT_CT_NEO at 43 days (range 28-63), and SX at 48 days (range 31-72 days). PHT_IOTT and RT recorded the longest median time to treatment initiation at 51 (range 34-89) and 53 days (range 27-85 days), respectively. The time to treatment associated with specific patient pathways is depicted in Figure 2.

Surgery treatment was provided to 19.64% of patients within 4 weeks, to 41% within 6 weeks, and later than 8 weeks to 40.5%. Systemic PHT was administered within 4 weeks to 23.62%, and RT to 25.5% of patients (Table 1 and online supplemental Figure S1).

**Table 1.** Cumulative distribution of first line treatment (FLT) since the initial bronchoscopy.

|  | Type of FLT | Pts who received FLT | Pts who did not received FLT |
| --- | --- | --- | --- |
| < 4 weeks | SX | 20% | 80% |
|  | RT | 26% | 74% |
|  | PHT | 24% | 76% |
| < 6 weeks | SX | 41% | 59% |
|  | RT | 39% | 61% |
|  | PHT | 47% | 53% |
| < 8 weeks | SX | 60% | 40% |
|  | RT | 52% | 48% |
|  | PHT | 66% | 33% |

*FLT* first line treatment; *PHT* pharmacotherapy; *RT* radiotherapy; *SX* surgery

### Treatment within COC

The majority, 85% (10,929 patients), of all treated patients received their first-line treatment within COCs (see online supplemental Table S1). The distribution of first-line treatments within COC varied across treatment modalities; practically all patients who received PHT_IOTT had their treatment in COC (reported 100%), followed closely by those who had SX (98%). Conversely, only 76% of patients receiving PTH_CT were administered within COCs.

There was a significant difference in overall survival (Log-rank p < 0.0001) between patients treated within COC and those treated elsewhere, with the former group being associated with better survival outcomes over time (Figure 3). This holds for patients receiving PHT_CT (Log-rank p< 0.0001) and RT (Log-rank p< 0.0001) as first-line treatments. However, for patients undergoing SX, the difference was not significant (Log-rank p= 0.45).

**Figure 3.**
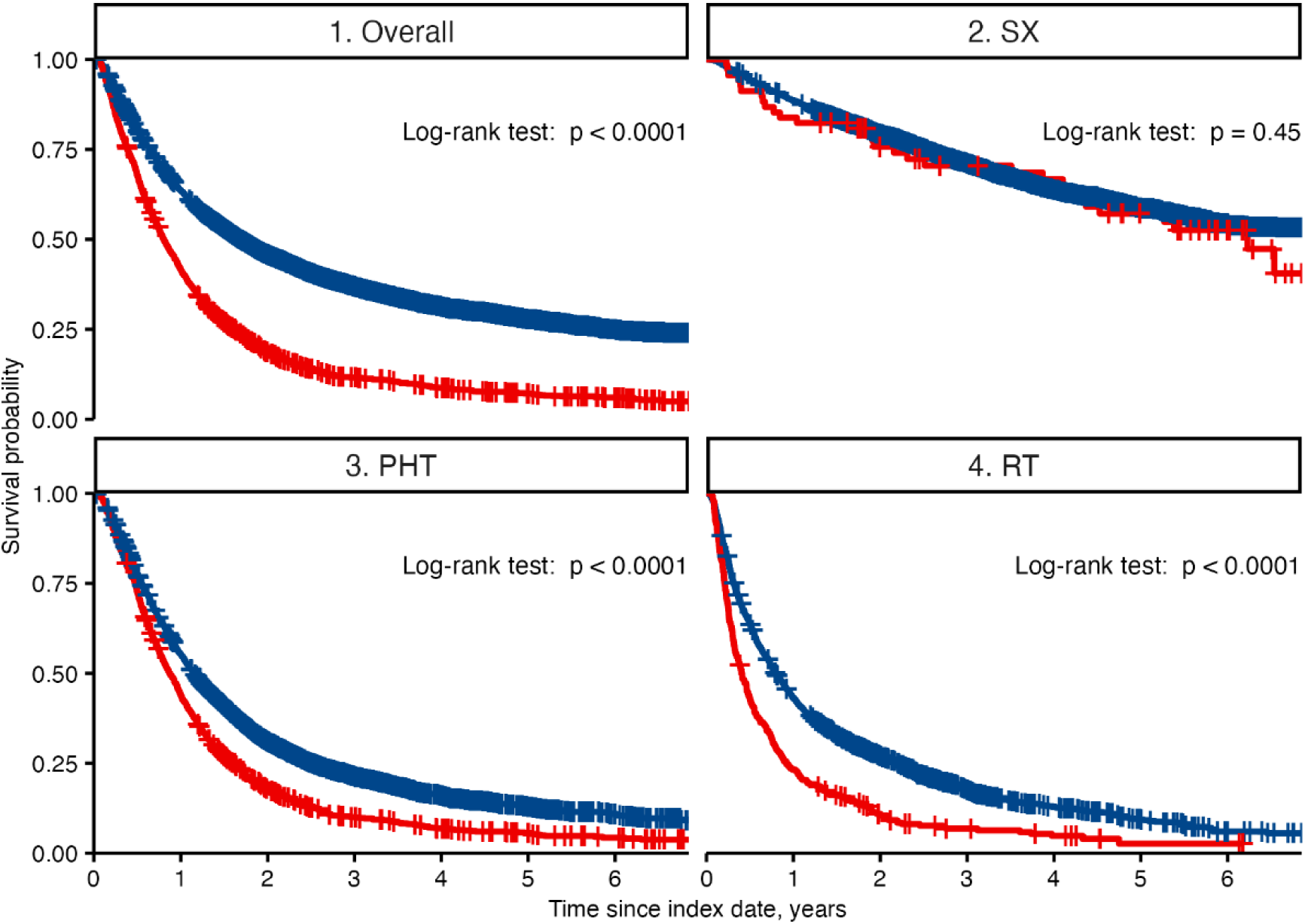
Comparative survival analysis of lung cancer patients treated within Complex Oncological Centers (COCs) or elsewhere, stratified by treatment modality. A Kaplan-Meier estimation with log-rank test (unadjusted) compares patients receiving first-line treatment within complex oncology centers (COCs) (blue line) versus those receiving first-line treatment outside complex oncology centers (COCs) (red line). Patients were treated with pharmacotherapy (PHT), including chemotherapy and precision therapy, surgery (SX), or radiotherapy (RT).

### Patient outcomes

A significant proportion of the incident cohort, 76% (12,691), had died by the end of the study period, with median overall survival of 339 days. Among those treated, the overall survival rate was 31% (see online supplementary Table S1), and the median survival was 533 days. Comparing the various modalities of first-line therapy, patients who underwent SX demonstrated the highest survival at 64.3% (median survival not reached). Patients with advanced disease treated with PHT_CT_NEO had a median survival of 1695 days, while those treated with RT had the lowest survival of 266 days (Figure 4).

**Figure 4.**
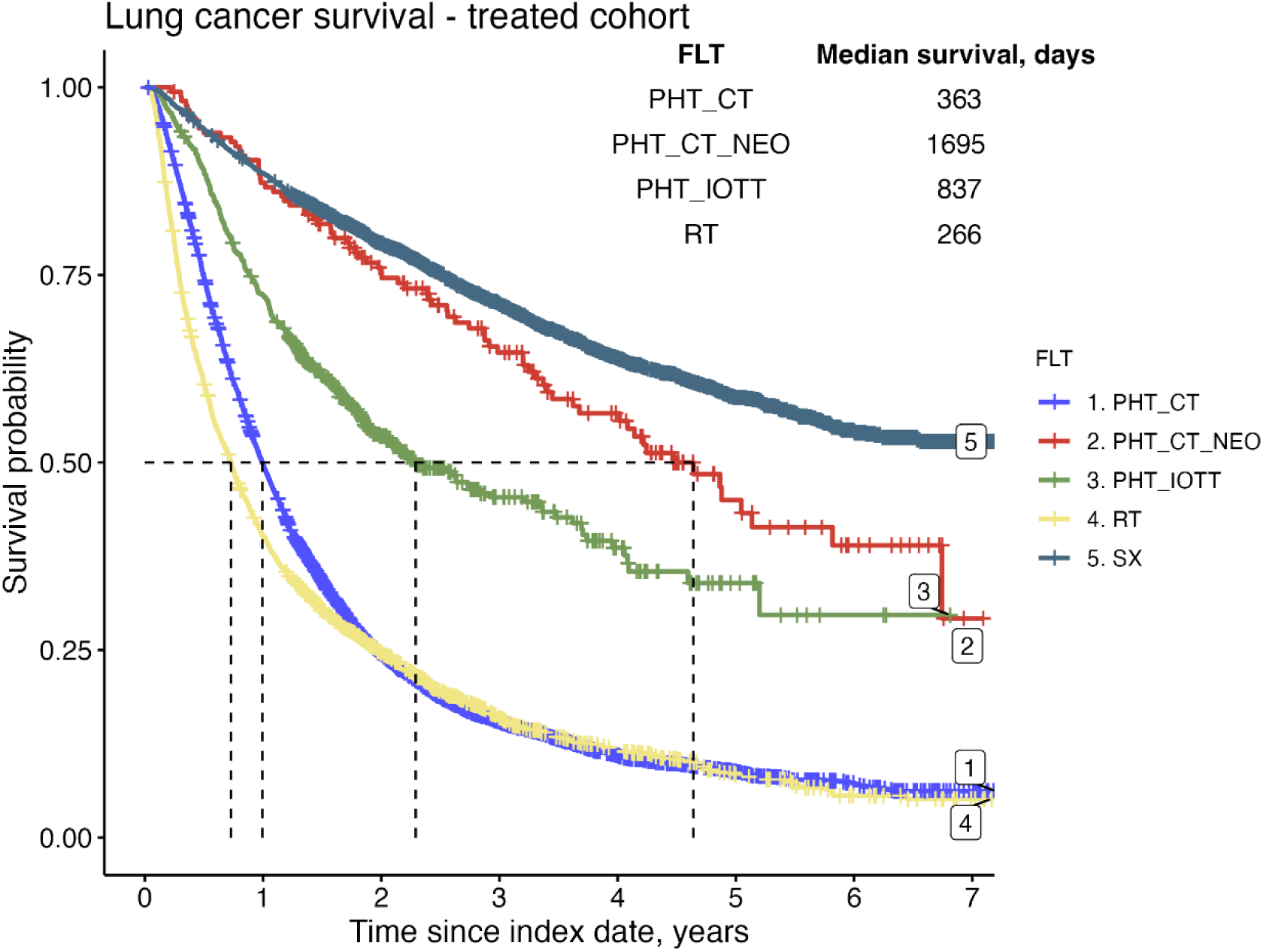
Comparative survival analysis of lung cancer patients by first-line treatment modality. Survival using a Kaplan-Meier estimation was analyzed from the initial bronchoscopy and stratified by the first-line treatment (FLT), including chemotherapy (PHT_CT), neoadjuvant chemotherapy (PHT_CT_NEO), precision therapy (PHT_IOTT) comprising immunotherapy (IO) and targeted therapy (TT), radiotherapy (RT), or surgery (SX).

Examining the survival outcomes across various treatment pathways for NSCLC patients, trajectories involving SX interventions exhibit notably favorable outcomes (see online supplemental Figure S2). Similarly, patients treated with PHT_CT_NEO achieved high 2-year survival rates compared to patients treated with systemic therapy for advanced disease. In general, trajectories with MDT demonstrate enhanced 2-year survival rates, typically ranging from a 7 to 14 percentage point increase over non-MDT pathways (see online supplemental Table S2).

## DISCUSSION

The period of the diagnostics as well as immediately following the identification of a lung cancer offers a critical window for effective intervention into the system offering the medical care. Our aim was to provide a clear depiction of the current state in the pathways of NSCLC patients, facilitating a foundation for potential improvements and assessing the impact of upcoming lung cancer screening. We developed a methodology for administrative data processing which can be implemented as a technical infrastructure to fulfill the organization and quality evaluation of cancer care in the Czech Republic. The comprehensive ICT model developed by Institute of Health Information and Statistics of the Czech Republic, integrating multiple data sources, has been prepared as a legal basis for the collection and publication of cancer care performance indicators. Patient pathway mapping is one of the key quality indicators that need to be implemented near the Czech National Cancer Registry (cancer notification, pathology result) to utilize administrative reimbursement data in the integrated system for the classification of cancer care.

In our work, we have described the individual points in the patient pathway that can be measured separately in different regions of the Czech Republic and/or at different time points, becoming effective keys for assessing access to care. The study was not designed with confirmatory intentions, nor was it aimed at establishing causality. This is primarily due to the absence of detailed staging information within the dataset, which inherently limits our ability to conclusively rule out selection bias or other confounding factors as explanations for the observed discrepancies. This is why we particularly discuss the importance of analyzing the patient pathway as a whole.

An optimal care pathway for lung cancer is still in its preliminary stages across broader health systems [27]. Significant variations exist in definitions of the intervals used to describe the timeliness of care for lung cancer [28]. Furthermore, there is little evidence about the effects of pathways used in oncological care [29]. Conversely, large electronic health records and administrative data have proven invaluable for understanding this subject. Integration of specific quality improvement measures is recommended by the European Respiratory Society in lung cancer care [30], and the general objective of the iPAAC [26] is to develop innovative approaches to advances in cancer control. Pilot 7 evaluates the feasibility of linking population-based cancer registry datasets with administrative and health data sources to describe the complete pathway of cancer patients and to assess the adherence of administered treatments to standard clinical guidelines.

With respect to the missing patient selection, in the discussion of patient trajectories, we will further focus only on time to treatment, access to COCs, and involvement of MDTs. Generally, decisions made during the critical period of disease diagnostics have the potential to significantly influence overall outcomes. The principle that cancer should be diagnosed swiftly, and promptly treated based on accumulated insights and expertise is widely regarded as historic axiom in oncology [17,31]. The pandemic had an impact on the availability of biopsies (BX) and the work of pulmonologists. Additionally, many COVID-19 symptoms overlapped with cancer symptoms, which may have led to late detection of lung cancer. As the time period of the analysis coincided significantly with the period of the COVID-19 pandemic, this overlap may have contributed to over-reporting of deaths. Therefore, it would be interesting to examine when and why those who didn’t receive treatment died, specifically. An analysis including the number of biopsies and the duration of diagnosis may serve as an indicator of the quality of centers performing NSCLC diagnosis. Moreover, an index date starting on the date of the first symptom of NSCLC or the first contact of the patient with a doctor who referred them for a BX would be a better index date, possibly demonstrating the diagnostic delay from the first symptom to the first BX.

Despite repeated attempts to determine the critical time for treatment initiation in lung cancer patients, consistent evidence is lacking [32,33], primarily due to methodological heterogeneity in defining the starting point for timing the intervention. A systematic review of European studies examined times to diagnosis and treatment of lung cancer and concluded that they are often longer than recommended [32]. Our data are consistent with those from Europe, with the median time to treatment ranging between 28-71 days in the Czech Republic, compared to the EU range of 30-84 days [32], which often exceeded published recommendations. Factors associated with timeliness have been incompletely examined, and it remains unclear whether more timely care improves outcomes. Our analysis provides a detailed overview of the time intervals between individual points in the patient’s journey.

We have shown that chemotherapy is associated with the shortest time to treatment initiation (42 days), while the duration for initiating precise therapy is one week longer, with a median of 51 days. This reduction could be due to the omission of extensive diagnostic procedures necessary for determining subsequent precision-based therapies, whether administered directly or after the first-line therapy. Furthermore, preoperative examinations or precise mapping of the affected area for radiation therapy are omitted from the process. It is possible that many of those with chemotherapy as the first-line therapy are prognostically worse patients excluded from indication for precision treatment due to poorer performance status. Time to precise therapy is critical for patients in the advanced stage of lung cancer and could be potentially measured as quality indicator in COCs.

A recent study suggests that a time to surgery longer than 4 weeks is associated with an increased rate of recurrence and death [34]. One of the reasons for the ambiguity in this area is the precise determination of the moment when the patient should be urgently directed towards further steps. There is an apparent paradox, where highly suspicious patients with well-manifested diseases will be promptly managed and referred for targeted examinations before treatment or directly to treatment. However, these patients will likely suffer from advanced stages of the disease, and the benefit of early intervention will likely result in only a mild extension of life. Conversely, for patients in early stages of the disease (Stages I and II), where early precise determination and subsequent treatment can lead to healing, the duration to treatment is longer due to uncertainty, new and repeated examinations, which apparently have a prognostically unfavorable impact. Czech data presented by our analysis are highly unsatisfactory because they show that only 20% of NSCLC patients undergo surgery within 4 weeks, and even within 8 weeks from diagnosis, the number of operated patients does not exceed 70%. However, the observed period includes the time before (2018-2019) and after (2020-2021) the centralization of surgical treatment in the Czech Republic, which may have an impact on the duration to surgery. Inclusion of disease stage is crucial for accurate monitoring of this indicator.

Additionally, it’s plausible that treatments, particularly those like chemotherapy that are more immediate and less reliant on extensive diagnostics, are commenced directly at the center where the diagnosis was made, avoiding patient delegation to COCs. Centralized care in high-volume centers ensures that patients benefit from the expertise and resources available in such settings, potentially leading to more accurate diagnoses and tailored treatment plans. Despite challenging methodology, emerging evidence suggests that centralized high-volume care, the involvement of MDT, and minimizing time to treatment initiation are pivotal factors that could enhance overall survival rates for NSCLC patients [20,21,32,34–35]. The most complex treatment is provided in a network of 15 COCs in the Czech Republic, 10 of which are certified lung cancer centers and five are highly specialized thoracic surgery cancer centers [4]. We have shown that lung cancer surgery and personalized treatment are predominantly centralized in our country which is good starting point for improving the quality of care. Our comparison of the prognosis of patients operated on in COC vs. elsewhere was limited by the fact that 98% of patients fell into the first group. Although the survival data of operated patients are in line with those published [10], interpreting them without staging data must be done with caution. Conversely, patients treated with systemic therapy and radiotherapy significantly benefited from centralized treatment. However, the challenge remains to ensure early access to both surgical and systemic care for NSCLC patients across the regions of the Czech Republic.

Most studies focused on the association between MDT and outcomes are, among other reasons, non-randomized and non-blinded for ethical considerations. The existing evidence does not unequivocally indicate an association between MDT and survival in NSCLC patients [18,19]. We showed a trend of better survival in trajectories involving an MDT review; however, caution must be exercised without considering the underlying patient selection processes. For example, patients with better performance status may be preferentially discussed in MDT meetings. It is particularly important that about half of operated patients do not undergo an MDT review. It is possible that in some patients who underwent MDT, this consultation was not documented. Although the reporting of the signal code for MDT, as we know it now, has been in effect since January 1, 2017, our data show that the reporting of MDT has been increasing over time. It is also worth mentioning that the data collection system for insurance companies, known as care reporting, is not standardized in the Czech Republic.

Screening of high-risk individuals, as recently initiated in the Czech Republic and elsewhere, may deliver load of early-stage patients with potentially curable stages [22,36]. While historical incidence rates for pulmonary nodules in the US remained steady for many decades at 150,000 per year [37], a more recent evaluation using electronic health records and natural language processing increased the incidence tenfold to over 1.5 million nodules detected per year [38]. This will be a substantial game-changer. The opportunity window for curable surgery in those early diagnosed demands a prompt and timely response to be effectively utilized. Conversely, the influx of new patients may quickly create a capacity bottleneck in the centers. Mapping the patient journey is very practical in this regard, highlighting the system’s limits in ensuring timely access to treatment or, conversely, in measuring the effectiveness of care centralization. Measuring time to treatment over time can serve as a quality indicator for benchmarking care-providing centers.

Our analysis has several limitations: it lacks information on disease stage, histopathological subtype, and causes of death. The index date starts from the date of biopsy examination rather than reflecting the onset of disease. Administrative claim reporting lacks robust standards.

## CONCLUSIONS

Mapping the patient pathway serves as a quality indicator. The integration of elements— centralized high-volume care, multidisciplinary approaches, and timely treatment—into NSCLC management protocols requires robust evidence to substantiate their effectiveness. The emphasis on these factors extends beyond clinical considerations to encompass economics and logistics, underscoring the importance of efficient healthcare delivery systems that optimize patient outcomes while considering cost-effectiveness. This evidence provides the foundation for developing guidelines and policies aimed at optimizing care delivery in high-volume centers, where such interventions are most feasible and likely to yield significant benefits for patients. As the healthcare community continues to strive for improved NSCLC survival rates, these factors should remain at the forefront of early intervention strategies, highlighting the need for ongoing research and evidence-based practice in this evolving field.

## SUPPLEMENTARY MATERIALS

### Additional file 1 (PDF format)

**Table S1** Characteristics of patients treated for lung cancer diagnosed from January 2017 through 2022

**Figure S1** Cumulative distribution of first line treatment (FLT) since the initial bronchoscopy

**Figure S2** Comparative survival analysis of lung cancer patients by pathway trajectory

**Table S2** Survival outcomes by patient pathway trajectory since index date

## DECLARATIONS

## Acknowledgments

The data were kindly provided by the Health Insurance Bureau in Prague, Czech Republic.

## Authors’ contributions

Conception/design: A.T., G.D., M.R. Investigation: G.D. Data analysis: A.T., G.D. Project administration: A.T. Data interpretation: A.T., G.D., Z.B., L.D., V.K., V.S., P.T. Manuscript writing: A.T., G.D., Z.B., K.M. Manuscript editing: L.D., V.K., V.S., P.T., J.M. Final approval of the manuscript: All authors.

## Ethics approval

This study was conducted using anonymized retrospective administrative claim data, and approval by an ethics committee was not required.

## Data availability

The data remains property of the insurance companies and cannot be shared without their permission.

