## Supplementary materials for "Treatment pathways of non-small cells lung cancer patients in the Czech Republic: an insight from administrative claims data"

### Supplementary material

Table S1 Characteristics of patients treated for NSCLC diagnosed from January 2017 through 2022

| | Overall<br>N = 12,803 | CT<br>N = 6,071 | IOTT<br>N = 681 | CT_NEO[3]<br>N = 166 | RT<br>N = 2,356 | SX<br>N = 3,529 |
|---|---|---|---|---|---|---|
| **Age** | 69 (63, 74) | 69 (63, 73) | 69 (60, 74) | 66 (61, 70) | 71 (65, 77) | 68 (62, 72) |
| **Males** | 8,053 (63%) | 4,093 (67%) | 358 (53%) | 111 (67%) | 1,510 (64%) | 1,981 (56%) |
| **Females** | 4,750 (37%) | 1,978 (33%) | 323 (47%) | 55 (33%) | 846 (36%) | 1,548 (44%) |
| **Survived[1]** | 3,954 (31%) | 869 (14%) | 350 (51%) | 90 (54%) | 377 (16%) | 2,268 (64%) |
| **Died[1]** | 8,849 (69%) | 5,202 (86%) | 331 (49%) | 76 (46%) | 1,979 (84%) | 1,261 (36%) |
| **MDT absent[2]** | 6,359 (50%) | 3,349 (55%) | 248 (36%) | 75 (45%) | 1,138 (48%) | 1,549 (44%) |
| **MDT reported[2]** | 6,444 (50%) | 2,722 (45%) | 433 (64%) | 91 (55%) | 1,218 (52%) | 1,980 (56%) |
| **FLT in COC** | 10,929 (85%) | 4,634 (76%) | 681 (100%) | 143 (86%) | 2,010 (85%) | 3,461 (98%) |
| **FLT not in COC** | 1,874 (15%) | 1,437 (24%) | 0 (0%) | 23 (14%) | 346 (15%) | 68 (1.9%) |
| **Median time to treatment** | 46 (28, 71) | 42 (28, 64) | 51 (34, 89) | 43 (28, 63) | 53 (27, 85) | 48 (31, 72) |
| **Time to treatment grouped** | | | | | | |
| **[0,30)** | 3,444 (27%) | 1,785 (29%) | 131 (19%) | 46 (28%) | 657 (28%) | 825 (23%) |
| **[30,60)** | 4,946 (39%) | 2,510 (41%) | 276 (41%) | 70 (42%) | 657 (28%) | 1,433 (41%) |
| **[60,120)** | 3,365 (26%) | 1,406 (23%) | 140 (21%) | 44 (27%) | 744 (32%) | 1,031 (29%) |
| **[120+)** | 1,048 (8.2%) | 370 (6.1%) | 134 (20%) | 6 (3.6%) | 298 (13%) | 240 (6.8%) |

[1]Within a period of 2017 through 2022. [2]Some MDT consultations could have been reported under different codes, such as individual claims for each healthcare provider specialist, or they might not have been reported at all. [3]CT prior surgery, following no later than 6 month after its onset.
PHT, pharmacotherapy; CT, chemotherapy; CT_NEO, neoadjuvant chemotherapy; IO, immunotherapy; TT, targeted therapy; BX, bronchoscopy; RT, radiotherapy; SX, surgery; MDT, multidisciplinary team; FLT, first-line therapy; COC, complex oncological center.

Figure S1 Cumulative distribution of first line treatment (FLT) since the initial bronchoscopy

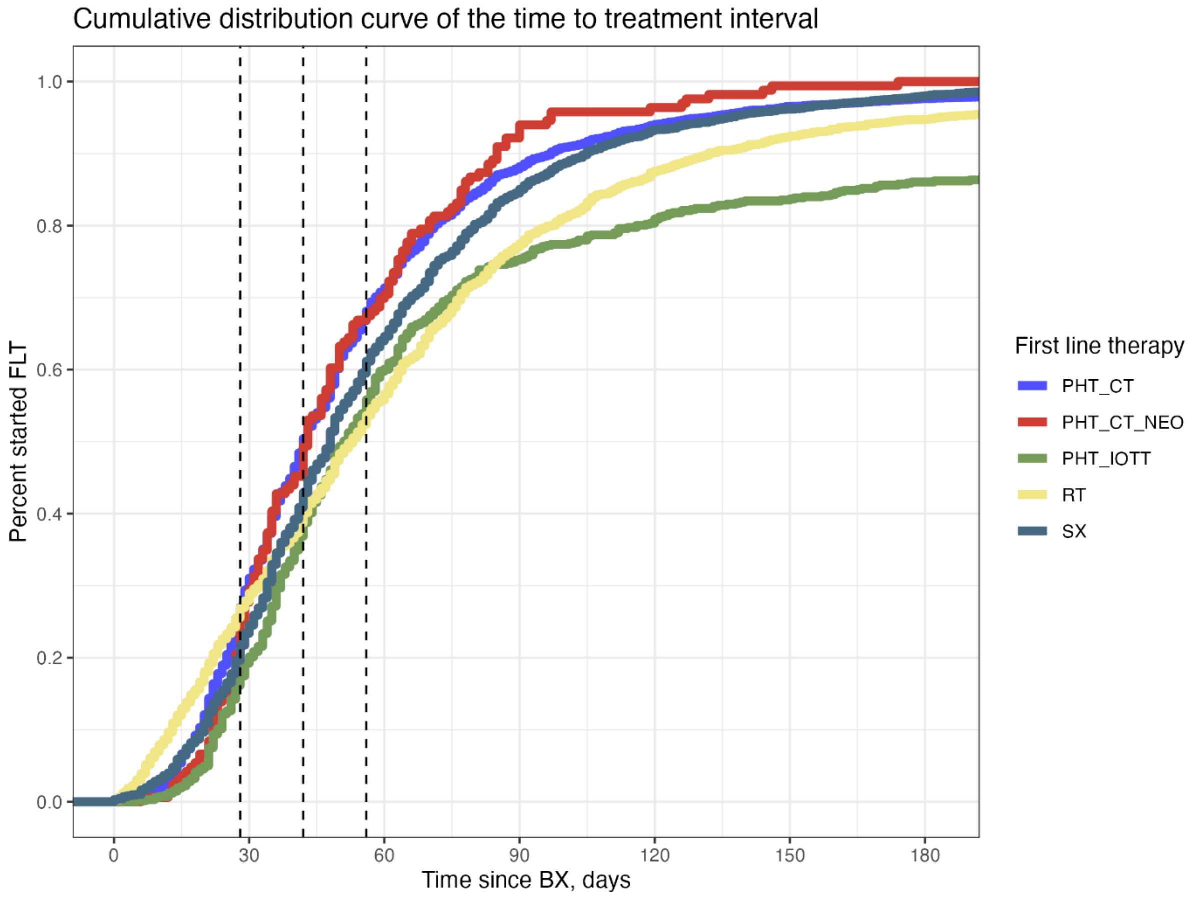


Patients receiving first-line treatment chemotherapy (PHT_CT), neoadjuvant chemotherapy (PHT_CT_NEO), precision therapy comprising immunotherapy and targeted therapy (PHT_IOTT), radiotherapy (RT), or surgery (SX) since the initial bronchoscopy. Dashed lines indicate the four, six and eight-week interval.

Figure S2 Comparative survival analysis of lung cancer patients by pathway trajectory

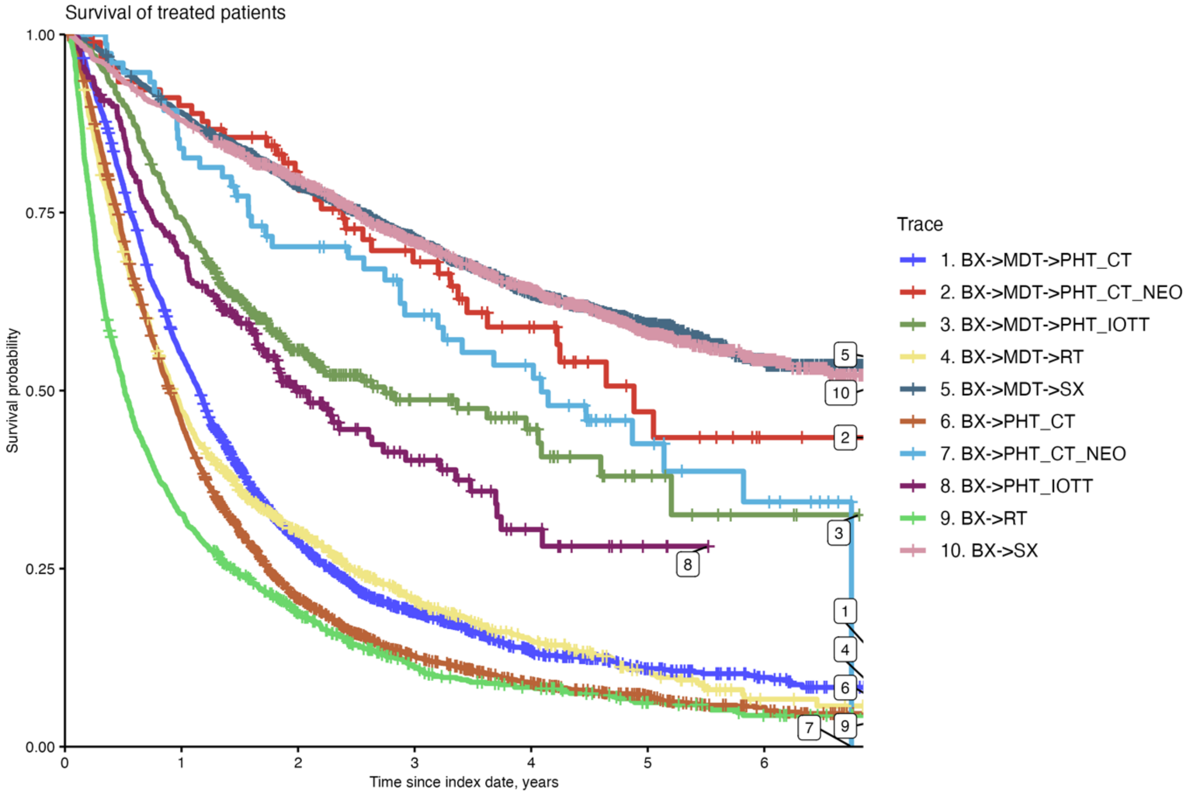


Survival using a Kaplan-Meier estimation was analyzed since the initial bronchoscopy (BX) and stratified by their trajectory to the first line treatment; chemotherapy (PHT_CT), neoadjuvant chemotherapy (PHT_CT_NEO), precision therapy comprising immunotherapy and targeted therapy (PHT_IOTT), radiotherapy (RT), or surgery (SX) either directly or via multidisciplinary team review (MDT).

Table S2 Survival outcomes by patient pathway trajectory since index date

| Trajectory | Median survival (Days) | Median Survival 95% CI | SR 90 Days | SR 180 Days | SR 365 Days | 2-Year SR |
|---|---|---|---|---|---|---|
| BX-SX | NA | 2273 - NA | 97% | 93% | 88% | 80% |
| BX-MDT-SX | NA | NA | 99% | 95% | 89% | 79% |
| BX-RT | 187 | 171-208 | 73% | 51% | 33% | 19% |
| BX-MDT-RT | 339 | 315-364 | 86% | 69% | 47% | 30% |
| BX-PHT | 348 | 336 - 362 | 89% | 73% | 48% | 24% |
| BX-MDT-PHT | 453 | 439 - 478 | 93% | 81% | 59% | 34% |
| *BX->MDT->PHT_CT_NEO* | 1782 | 1323 - NA | 99% | 93% | 90% | 81% |
| *BX->PHT_CT_NEO* | 1492 | 1168 - NA | 100% | 95% | 84% | 70% |
| *BX->PHT_CT* | 329 | 316 - 343 | 88% | 71% | 45% | 21% |
| *BX->MDT->PHT_CT* | 419 | 400 - 436 | 93% | 79% | 55% | 29% |
| *BX->PHT_IOTT* | 725 | 607 - 997 | 94% | 86% | 69% | 50% |
| *BX->MDT->PHT_IOTT* | 1000 | 759 - 1492 | 97% | 90% | 74% | 56% |

The pathway trajectories are categorized based on the involvement of bronchoscopy (BX), multidisciplinary team consultations (MDT), pharmacotherapy (PHT), radiotherapy (RT), and surgical interventions (SX). Among pharmacotherapies, chemotherapy (PHT_CT), precision therapy (PHT_IOTT), and neoadjuvant chemotherapy (PHT_CT_NEO) were further defined. The 95% confidence intervals (CI), and survival rates (SR) at various time points post-treatment initiation are provided. "NA" indicates data not available or not applicable. The index date was operationalized as the date of the first BX.
